# The effect of non-genetic determinants of human milk oligosaccharide profiles in milk of Ugandan mothers

**DOI:** 10.1101/2024.03.21.24304661

**Authors:** Tonny Jimmy Owalla, Victor Irungu Mwangi, Sara Moukarzel, Emmanuel Okurut, Chloe Yonemitsu, Lars Bode, Thomas G. Egwang

**Author notes:** **Corresponding author**: Tonny Jimmy Owalla, P.O. Box 9364 Kampala-Uganda. **TJO current address:** Department of Global Health, University of Washington, Seattle, WA, USA.

## Abstract

**Background:** Human milk oligosaccharides (HMOs) protect against infection and promote growth and cognitive development in breastfeeding children. Non-genetic factors which influence HMO composition in breastfeeding mothers in rural Africa have not been investigated.

**Objective:** We undertook a cross-sectional study to determine the association between HMO profiles and non-genetic maternal factors and children’s sex in Ugandan mother-children pairs.

**Method:** Human milk was collected from 127 breastfeeding mothers by manual expression. HMO analysis was by high performance liquid chromatography. The proportion of each HMO per total HMO concentration was calculated. Spearman’s correlation and Mann-Whitney U test were used to assess the relationship between individual HMOs and maternal factors and infant sex.

**Result:** Nineteen HMOs were assayed. The prevalence of secretor and non-secretor status, based on the proportion of mothers with high milk concentrations of 2’FL and LNFP 1, was 80.3 % and 19.7 %, respectively. In secretor mothers, 2’FL, DFLac and LNFP I constituted > 57 % while in non-secretor mothers LNT and LNFPII constituted 46.9 % of the measured total HMOs. The median 3’SL concentration in milk of all mothers of male children was significantly higher than that in all mothers of female children. The median DFLac concentration in all mothers was significantly higher in multiparous mothers compared to primiparous mothers. Higher FDSLNH and lower LNH concentrations were observed in overweight secretor and non-secretor mothers, respectively. Median concentrations of LNFP I and DSLNT were significantly higher in all mothers < 18 years old compared all mothers > 18 years old. Concentrations of specific HMOs increased, decreased, or remained unchanged with increasing lactation duration in secretor and non-secretor mothers.

**Conclusions:** Specific HMOs were associated with infant sex and maternal age, parity and post-partum BMI in Ugandan mothers but were different from those reported in other populations.

## Introduction

Human milk oligosaccharides (HMOs), comprises of over 150 non-digestible unconjugated glycans and constitutes the third most abundant solid component in human milk-besides lipids and fats (1). HMOs play a critical role in protecting infants against infection through modulation of gut microbiota and the immune system (2–6). These HMO effects are composition- and structure-specific (3, 6). The HMOs basic structure consist of a lactose core which is modified by glycosyltransferases to form 4 main types of HMOs, namely, non-fucosylated neutral HMOs containing N-acetylglucosamine at the terminal end, neutral fucosylated HMO containing fucose at the terminal position, sialylated HMOs containing sialic acid, and HMOs containing both fucose and sialic acid (1,7,8). HMO composition and concentration are influenced by maternal genetics. Breastfeeding mothers with an active gene encoding the α (1,2)-fucosyltransferase (FUT2) enzyme have milk with characteristically high levels of 2’-FL and Lacto-N-fucopentaose I (LNFPI) and are designated secretor mothers (6). Women who do not express FUT2 have milk lacking these HMOs and are designated non-secretors. The prevalence of secretor mothers vary by geographic from a high prevalence in South America to the lowest prevalence in African countries ranging from 63% in South Africa to 75 % in Malawi and Kenya (9).

There is evidence that environmental, non-genetic maternal and infants’ factors are associated with differential HMO concentration in Asian, European, Canadian and American populations (9,10, 11). However, there is a dearth of information about the non-genetic factors that influence HMO composition and concentrations in breastfeeding mothers in African rural settings. Our study investigated for the first time non-genetic maternal and infant factors that influence HMO composition and concentration in Ugandan breastfeeding mother-child pairs in rural northeastern Uganda.

## Methods

### Study population and design

The study population was part of a cohort who were participating in a prospective longitudinal study investigating the effectiveness of a malaria vector control intervention in Abwokodia Parish, Katakwi district in Northeastern Uganda. Details of the study site have been reported elsewhere (12). The breast milk sub-study was a clinic based cross-sectional survey of breastfeeding mother-infant pairs. All breastfeeding infants less than two years old and their respective breastfeeding mothers were considered eligible for the study upon consent by one or both parents as applicable. The breastfeeding infants were part of 400 children under 5 years old who were being followed up for malaria prevalence and incidence. All the identified 127 mother-infant pairs were included in the study (**Supplementary Figure 1**).

### Participants recruitment and anthropometric measurements

The study was conducted in March 2018. Mothers of all breastfeeding infants participating in the longitudinal study were mobilized by a social scientist with the help of village health teams (VHTs). Mother-infant pairs visited the study clinic at St. Anne Health Center III, Katakwi district for screening and enrolment. The objectives of the study were explained to the mothers and informed consent obtained. Human milk samples were collected from consenting mothers. The demographic characteristics (infant sex and maternal age, parity, BMI, lactation duration), and other anthropometric data for mother-child pairs were recorded on a standardized questionnaire.

### Breast milk sampling and treatment

A single 5-mL human milk sample was collected from each of one hundred and twenty-seven lactating mothers using manual expression with the help of a senior midwife who is also an experienced lactation nurse. Milk samples were collected into sterile 50-mL Falcon tubes and immediately frozen in dry ice (approximately −70°C). Milk samples were transported in dry ice to Med Biotech Laboratories headquarter in Kampala before being air-freighted in dry ice to the University of California San Diego for HMO high performance liquid chromatography (HPLC) analysis.

### HMO Extraction, Analysis, and Secretor Status Determination

HMO (the 19 well characterized and most abundant) analysis was performed at the University of California, San Diego, as previously described (13), by using HPLC after fluorescent derivatization. Raffinose was added to each milk sample as an internal standard for absolute quantification. The total concentration of HMOs was calculated as the sum of the specific oligosaccharides detected. The proportion of each HMO per total HMO concentration was calculated. Maternal phenotypic secretor status was determined by the relative abundance (secretor) or near absence (non-secretor) of the (α 1-2) linked Fuc (2′(-fucosyllactose (2′FL) in the respective milk samples.

### Statistical Analyses

Statistical analyses were performed using GraphPad Prism version 9.0.2 and STATA version 15. The Shapiro Wilk test were used to evaluate the variables’ distribution including the oligosaccharide concentrations. Normally distributed data were compared using Student’s t-test, the t-test for paired samples or one-way analysis of variance for groups, while non-parametric comparisons were made using the Mann-Whitney U test for paired samples or Kruskal-Wallis test for groups. The one-way ANOVA and Kruskal-Wallis tests were followed up by the appropriate post hoc multiple comparison test. Chi-square or Fisher exact tests were also performed to compare categorical variables. Exploratory analysis were performed to compare median oligosaccharide concentrations according to infant sex, maternal age (years), current BMI (kg/m^2^), maternal parity (primiparous/multiparous), and duration of lactation (weeks). The results were presented as box-plot graphs. Correlations of the oligosaccharide concentrations with other infant or maternal variables were assessed using the Spearman rank test with 95% confidence interval. Significance was defined as *P* values of less than .05.

### Ethical consideration

The Uganda National Council for Science and Technology and the Research Ethics Committee of the Vector Division, Ministry of Health, approved the original HD4MC study. All mothers who donated milk samples and brought infants to the malaria clinic signed or thumb-printed an informed consent form.

## Results

### Characteristics of the Study Population

The study population comprised of 127 mothers and their breastfeeding children in Katakwi District in northeastern Uganda. The mothers had a mean age of 26.6 years (range 15-46) with a post-partum mean BMI of 23.3 kg/m2 (**Table 1**). Most of the mothers had had several children (> 78 %) and had breastfed for a mean of 41.3 weeks (range 3-103). The children had a mean age of 46.5 weeks (range 3-110) and 52.6 % were females. The mean birth weight was 3.1 kg (range 1-4.8). The mean children’s heights/lengths and weights at the time of sampling were 70.5 cm (range 50-98) and 8.80 kg (5-16), respectively. The mean mid-upper arm circumference for the children was 14.8 cm (range 12.1-18.7). The mean hemoglobin level in children was 10.9 g/dL (range 8-11.5). The children had a mean temperature of 36.5 °C (range 35.4-39.4) and the majority (97.7 %) had normal temperature at the time of sampling.

**TABLE 1:** Characteristics of the study population.

| **Characteristic** | **Mean** | **Range** | **Median ( IQR)** |
| --- | --- | --- | --- |
| **Mothers** |  |  |  |
| Age (years) | 26.57 | 15 - 46 | 25 (21 - 32) |
| *Postpartum BMI (Kg/m<sup>2</sup>) | 23.33 | 18.7 - 31.8 | 23.1 (21.9 – 24.4) |
| Lactation duration (Weeks) | 41.28 | 1 - 104 | 41 (23 - 60) |
| **Infants and young children** |  |  |  |
| Age (weeks) | 46.47 | 3 - 110 | 44 (28-66) |
| Birth weight (Kg) | 3.14 | 1 - 4.8 | 3.1 (2.8 - 3.5) |
| *Weight (Kg) | 8.80 | 5 - 16 | 9 (8-10) |
| *Height/length (cm) | 70.50 | 50 - 98 | 70 (64 - 76) |
| *Mid upper arm circumference (cm) | 14.82 | 12.1 - 18.7 | 14.7 (14-15.5) |
| *Temperature (°C) | 36.5 | 35.4 - 39.4 | 36.4 (36.2 – 36.7) |
| *Hemoglobin (g/dL) | 10.92 | 8 - 13.5 | 11 (10.3 - 11.8) |

\* = maternal and infant/child factors measured at sampling; Kg/m<sup>2</sup> = kilogram per meter squared; % = percent; cm = centimetre; °C = degree Celsius; g/dL = grams per decilitre; IQR = interquartile range; N = number of independent observations.

### HMO profiles and prevalence of secretor and non-secretor mothers

Mothers with an active secretor (Se) gene that encodes α (1,2)-fucosyltransferase are classified as “secretors” and their milk contains significantly higher concentrations of α(1,2)-fucosylated HMOs such as 2′FL, DFLac and LNFP I (6). The prevalence of Ugandan secretor and non-secretor mothers, based on the proportion of mothers with relatively high milk concentrations of 2’FL and LNFP 1 was 80.3 and 19.7 %, respectively (Table 1). In secretor mothers, 2’FL, DFLac and LNFP I constituted > 57 % of the measured total HMOs (**Supplementary Table 1**). By contrast, 2’FL, DFLac and LNFP I constituted only 1.9 % of the measured HMOs in non-secretor mothers in whom LNFP II and LNT alone constituted 46.9 % of the measured total HMOs.

### Infant sex is associated with differential concentrations of 3’SL in all mothers

The median 3’SL concentration in the milk of all mothers combined of male infants was significantly higher than those in milk of all mothers combined of female infants (701.8 (454.5-1059.8) nmol/mL versus 516.6 (309.8-853.8) nmol/mL; P =0.035) by the Mann-Whitney U test (**Supplementary Table 2**).

### Lactation duration is associated with differential HMO composition

Breastfeeding for 24 months or more is common in the study population and it was of interest to investigate HMO profiles beyond 12 months post-partum. We used Spearman’s correlation analysis to assess the relationship between HMO concentrations and lactation duration defined as the time elapsed from the time of first breastfeeding at birth to the time post-partum of breast milk sampling. Five and 10 out of the 19 HMOs had a significant positive and negative correlation, respectively, with lactation duration depending on mothers’ secretor status (**Supplementary Table 3**). The results of Mann-Whitney U tests comparing median HMO concentrations between mothers at different lactation durations closely paralleled those of correlation analyses. The concentrations of 3FL, 3’SL and DFLac were significantly higher in all mothers combined at > 6 months lactation compared to ≤ 6 months lactation (**Table 2**). By contrast, the concentrations of LNnT, 6’SL, LSTc, LNH, FLNH, DFLNH, and DSLNH were significantly lower in all mothers at > 6 months lactation duration. When the mothers’ secretor status was considered, LNFPI, LNnT, 6’SL, LSTc, FLNH, DFLNH, and DSLNH concentrations were significantly lower and 3 FL, 3’SL and DFLac concentrations were significantly higher in secretor mothers at > 6 months lactation (**Figure 1**). Similar HMO profiles were observed in non-secretor mothers but LNH and FDSLNH concentrations were significantly lower while LNFP I, FLNH and LNnT concentrations were unchanged in mothers at > 6 months lactation (**Supplementary Figure 2**). To see if the dynamics of HMO composition and concentrations persisted beyond 12 months (52 weeks), we compared HMO concentrations in groups of all mothers combined at lactation durations of ≤ 12 (N=21), 12-24 (N=14), 25-48 (N=40), 49-72 (36) and > 73 (N=16) weeks, respectively. The same HMOs increased, decreased, or remained unchanged, respectively, throughout lactation beyond 52 weeks (**Figure 2**). It was not possible to assess HMO profiles in similar groups of secretor and non-secretor mothers due to the small sample sizes.

**FIGURE 1.**
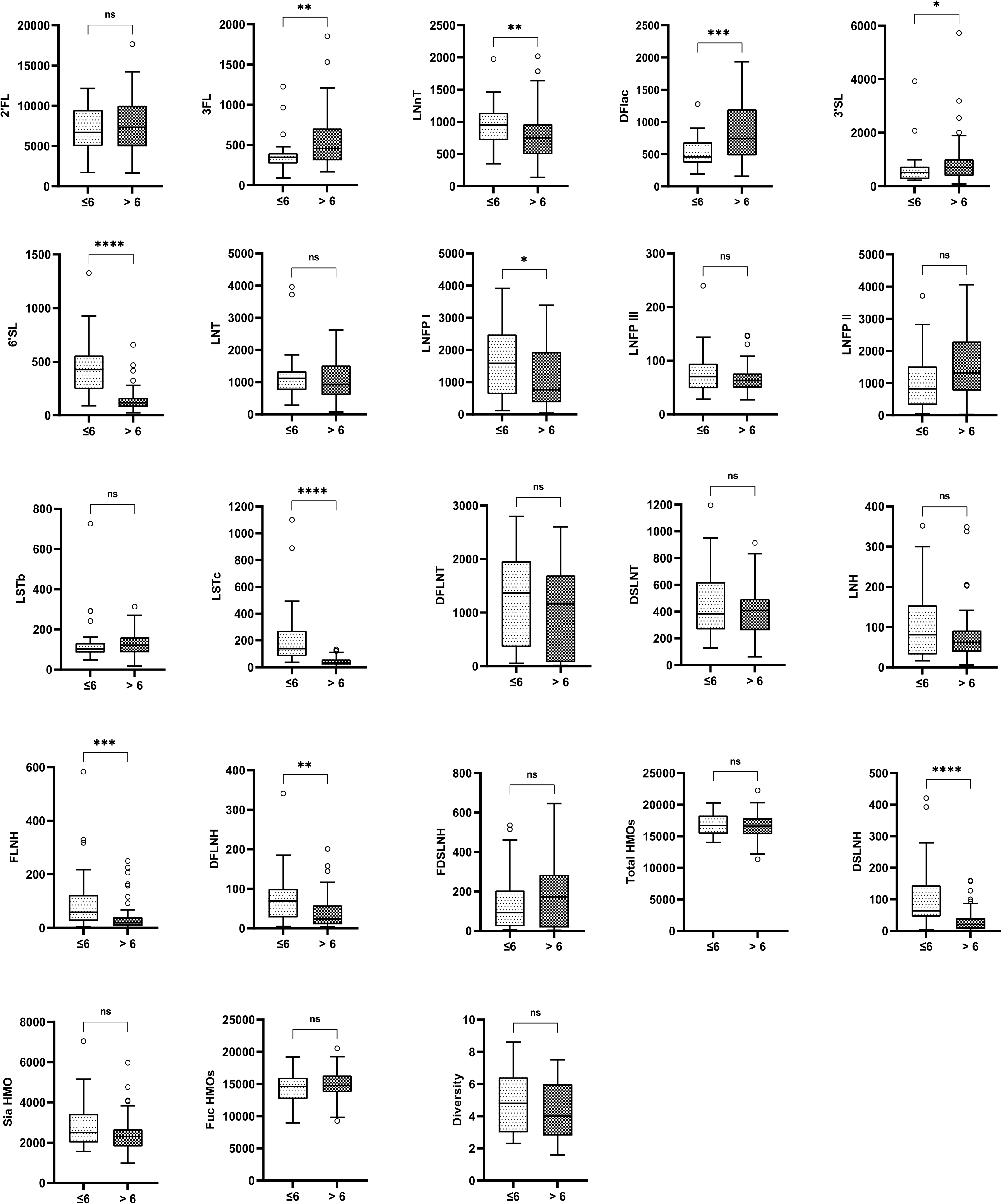
Concentrations of all measured HMOs before and after 6 months postpartum in breast milk from secretor Ugandan mother-child pairs. HMO name abbreviations are indicated in the list of abbreviations. Data are shown as box and whisker plots with the medians and the lower and upper quartiles or interquartile range (IQR). P values represent the outcomes of comparisons of two groups by the Mann-Whitney U test.

**FIGURE 2.**
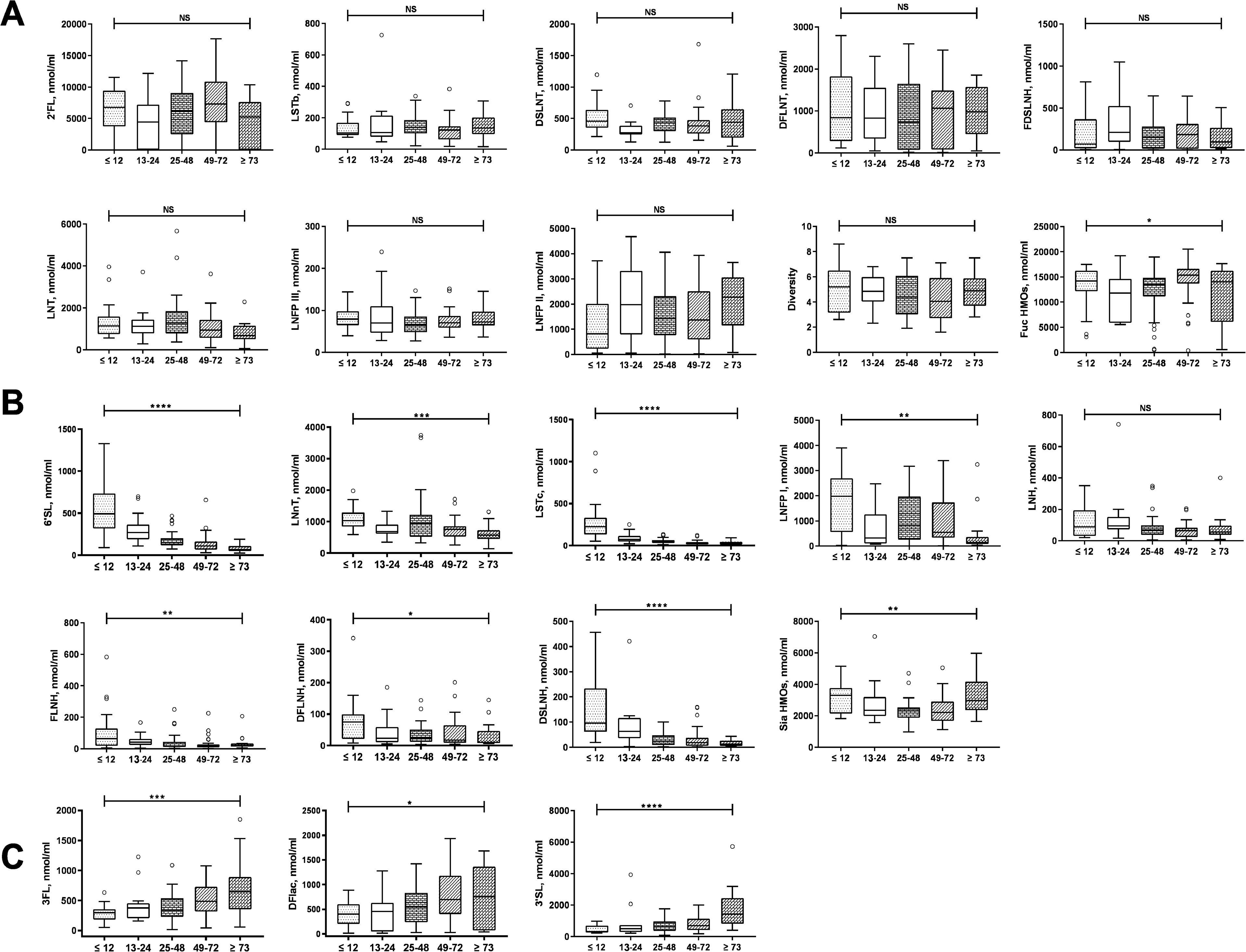
Concentrations of all measured HMOs in five groups of mothers representing different lactation stages ≤ 12 (N = 21), 13-24 (N= 14), 25-48 (N= 40), 49-72 (N= 36), ≥73 (N= 16) weeks postpartum in breast milk from Ugandan mother-child pairs. **A.** HMOs whose concentrations do not change from ≤ 12 to ≥73 weeks. **B**. HMOs whose concentrations decrease from ≤ 12 to ≥73 weeks. **C.** HMOs whose concentrations increase from ≤ 12 to ≥73 weeks. HMO name abbreviations are indicated in the list of abbreviations. Data are shown as box and whisker plots with the medians and the lower and upper quartiles or interquartile range (IQR). P values represent the outcomes of comparisons of two groups by the Mann-Whitney U test.

**TABLE 2:** Median (IQR) HMO concentrations and lactation duration in all mothers.

| HMOs<br>nmol/mL | Total (N=127) | >6 months (n=89) | ≤ 6 months (n=38) | Unadjusted<br>p-value |
| --- | --- | --- | --- | --- |
| 2'FL | 5929.5 (2715.8-9271.8) | 6379.9 (3640.8-9364.7) | 5189.9 (523.6-9044.3) | 0.21 |
| **3'FL** | 378.9 (263.8-606.8) | 452.3 (289.4-687.0) | 308.6 (209.2-400.8) | **0.002** |
| **LNnT** | 805.2 (578.2-1033.5) | 751.7 (506.9-977.6) | 937.7 (672.9-1140.3) | **0.020** |
| **3'SL** | 632.0 (353.4-987.8) | 731.6 (411.5-1101.7) | 470.0 (281.5-737.8) | **0.002** |
| **DFLac** | 548.4 (287.0-881.5) | 651.2 (378.9-1128.3) | 403.7 (192.0-594.8) | **<0.001** |
| **6'SL** | 157.8 (102.2-270.9) | 121.1 (82.3-171.6) | 403.4 (218.4-569.7) | **<0.001** |
| LNT | 1028.7 (694.8-1618.0) | 964.6 (646.1-1618.2) | 1134.7 (752.7-1366.6) | 0.42 |
| LNFP I | 599.3 (172.6-1863.9) | 493.7 (236.9-1707.5) | 1050.3 (109.3-2411.5) | 0.38 |
| LNFP II | 1411.1 (721.0-2551.2) | 1428.2 (782.5-2492.5) | 1218.7 (557.0-2796.8) | 0.89 |
| LNFP III | 70.0 (55.0-88.8) | 69.0 (57.1-83.9) | 79.0 (52.7-96.7) | 0.18 |
| LSTb | 123.5 (90.8-182.2) | 124.7 (94.8-181.9) | 114.2 (90.8-209.9) | 0.91 |
| **LSTc** | 46.5 (24.3-93.3) | 33.8 (19.3-57.5) | 133.1 (73.1-242.4) | **<0.001** |
| DFLNT | 865.9 (122.3-1576.6) | 904.5 (80.4-1481.6) | 825.8 (414.1-1677.1) | 0.13 |
| **LNH** | 71.2 (37.7-106.4) | 61.5 (37.2-90.8) | 86.1 (52.5-162.0) | **0.004** |
| DSLNT | 417.2 (274.3-512.6) | 426.5 (274.3-507.4) | 380.8 (280.5-512.6) | 0.98 |
| **FLNH** | 24.2 (13.2-57.0) | 20.5 (11.9-35.6) | 45.9 (19.7-101.2) | **<0.001** |
| **DFLNH** | 24.4 (9.4-66.1) | 20.3 (9.0-50.9) | 32.6 (14.5-80.4) | **0.030** |
| FDSLNH | 146.7 (21.1-310.1) | 168.4 (16.9-290.3) | 108.2 (30.6-450.2) | 0.27 |
| **DSLNH** | 31.7 (11.6-64.5) | 20.7 (6.9-38.5) | 81.8 (47.0-125.6) | **<0.001** |
| Total HMO | 16050.6 (13219.4-17668.9) | 16162.0 (13539.2-17668.9) | 15434.5 (11463.8-17416.4) | 0.74 |
| **Sia HMO** | 2395.0 (1995.9-3149.7) | 2347.5 (1926.0-2839.5) | 2766.7 (2062.5-3753.1) | **0.030** |
| Fuc HMO | 13962.3 (10903.9-15966.1) | 14074.6 (11403.9-16138.5) | 13461.9 (8989.9-14838.5) | 0.16 |
| Diversity | 4.7 (3.1-6.0) | 4.5 (3.0-5.9) | 5.2 (3.4-6.1) | 0.16 |

IQR = interquartile range; HMOs = human milk oligosaccharides; N = total number of mothers sampled; n = number of independent observations per category; > = greater than; ≤ less than or equal to. HMOs with significant unadjusted p values are in bold.

### Maternal Age is associated with differential concentrations of 2’FL, DFLac and DSLNT

2’FL and DFLac concentrations had a significant positive correlation with maternal age only in non-secretor mothers (**Supplementary Table 4**). LNnT had a significant negative correlation with maternal age in all mothers and secretor mothers but not non-secretor mothers. Interestingly, DSLNT concentrations had a significant negative correlation with maternal age in secretor mothers but a positive correlation in non-secretor mothers. Median concentrations of LNFP I and DSLNT were significantly higher in all mothers < 18 years old (N=4) than in all mothers > 18 years old (N= 123) by Mann-Whitney U test (**Supplementary Table 5**). It was not possible to compare median HMO concentrations between the two age groups by secretor status and lactation duration because of the small sample size for the younger mothers.

### Maternal Parity is associated with differential concentrations of DFLac in all mothers

To confirm if maternal parity affected HMO concentrations, we defined parity as primiparous mothers (N= 27) who have had one child versus multiparous mothers (N= 100) who have had more than one child. The median DFLac concentration was significantly higher in multiparous all combined mothers compared to primiparous all combined mothers by the Mann-Whitney U test (**Supplementary Table 6**). It was not possible to compare median HMO concentrations between both parity groups by secretor status and lactation stage because of the small sample size for primiparous mothers.

### Maternal BMI is associated with differential concentrations of FDSLNH and LNH

FDSLNH and LNH concentrations positively and negatively correlated with increasing maternal postpartum BMI at the time of sampling in secretor and non-secretor mothers, respectively (**Supplementary Table 7**). None of the other HMOs had any correlation with maternal BMI. When secretor mothers were classified into normal weight (BMI= 18.5-24.9, N= 81) and overweight (BMI = ≥25, N= 19) categories, median FDSLNH and LNH concentrations were significantly higher and lower, respectively, in overweight mothers by comparison with normal weight mothers by the Mann-Whitney U test in secretor and non-secretor mothers, respectively (**Supplementary Tables 8 and 9**).

## Discussion

Non-genetic factors including infant sex and lactation duration which influence HMO composition have been investigated in breastfeeding mothers from Brazil, China, Europe, and USA (9,11,15,16,17,19, 20,21,22). The majority of studies of the effect of lactation duration on HMO composition investigated limited lactation durations of 3-6 months; only one longitudinal study followed up breastfeeding mothers for 24 months (35). Despite the burgeoning literature on HMOs in breastfeeding mothers in African countries (9,33,35,36, 37, 38), there is a knowledge gap about non-genetic factors which influence HMO composition in mothers in rural and urban African settings. The aim of this study was to determine HMO profiles in 127 rural Ugandan mothers and assess their relationship with various maternal and infant factors. HMO profiles in Ugandan mothers demonstrated a high prevalence of secretors (80.3 %) with 2’FL and LNFP I predominating in secretors while LNFP II and LNT predominated in non-secretors (prevalence 19.7 %). Lower secretor prevalence has been reported in mothers from some African populations (9).

Infant sex and maternal parity, age, post-partum BMI and lactation duration were associated with specific HMO profiles in Ugandan mothers. First, higher concentrations of 3’SL were observed in mothers of male but not female children in this study. By contrast, in Brazilian and Chinese mothers significantly higher concentrations of LNT were associated with having male infants (15, 16). The fact that infant/child male sex is significantly associated with high concentrations of LNT and 3’SL in different studies involving different ethnicities suggests important differential roles for these HMOs in the health and development of male and female infants in the respective populations. Second, DFLac was significantly higher in milk of multiparous Ugandan mothers by comparison with primiparous mothers. Maternal parity has been associated with increased or decreased concentrations of LNT, LNnT and 3FL in European, Chinese, and Brazilian mothers (11, 15, 17). Third, LNFPI and DSLNT concentrations were significantly higher in Ugandan mothers under 18 years old by comparison with older mothers. In a single study involving a limited number of mothers of different ethnicities, maternal age was associated with increased or increased concentrations of specific HMOs which were different from those identified in this study (9). Finally, postpartum BMI was associated with higher and lower concentrations of FDSLNH and LNH, respectively, in Ugandan mothers. The relationship between pre-partum BMI and HMO composition is controversial (9, 15, 18, 19, 20,21,22). In this population of Ugandan mothers, the above maternal and infant factors were associated with specific HMOs which are different from those reported in other studies (9, 11,15,16,17,18–22); this observation underscores the importance of population context-specific HMO data. The mechanisms underlying the associations between specific HMOs with maternal factors and infant sex are not known (10).

Lactation duration was an important non-genetic maternal determinant of HMO profiles in Ugandan mothers. The fact that concentrations of the same HMOs decreased, increased, or remained unchanged with increasing lactation duration beyond 52 weeks in Ugandan mothers probably highlights their importance in the survival and development of breastfeeding infants and young children especially in resource-poor rural and urban Uganda where infant morbidity, mortality and malnutrition remains disproportionately high (23, 24, 25). In this study population, two HMOs, 2’FL and LNFPI, are associated with protection against malaria in breastfeeding children (Mwangi et al under review). In prior literature, 2’FL has been implicated in multiple functions including protection against infections, reduction of morbidity and use of antipyretics and antibiotics (26, 27); stimulation of brain development (28, 29); weight gain (19); and improvement of cognitive functions (30). 2’FL also promotes the selective growth of bifidobacteria and therefore probably influences the composition of gut microbiota (31) and microbiota-induced immune functions (32) in breastfeeding infants. 2’FL and 3’FL, which also increased during lactation in Ugandan mothers, have been implicated in the prevention of mortality in uninfected Zambian infants born to HIV-positive mothers (33). The exact mechanisms by which HMOs reduce morbidity, prevent mortality and promote growth and development are unknown but could be associated with their reported anti-infective activities and effects on immunity (31,32, 34). The mechanisms underlying differences in HMO profiles during lactation remain speculative (10). There is need for research on the regulation of the enzymes involved in HMO biosynthesis during lactation in Africa, a region fraught with the constant threat of malnutrition and food insecurity which affect HMO concentrations (39).

This study has several limitations. First, cause-effect relationships between non-genetic maternal and infant factors cannot be inferred due to the cross-sectional nature of this study. Second, due to the small sample sizes, we were unable to adjust for multiple testing in statistical analyses nor make direct comparisons between secretor and non-secretor mothers. This limitation will be addressed in a planned powered longitudinal study of breastfeeding mother-child pairs. Third, milk samples were collected at a single time point, rather than longitudinally over several time points during lactation to monitor postpartum changes in HMO profiles over time. However, this limitation is mitigated by the fact that our cross-sectional milk sampling from mothers representing a wide range of lactation stages produced HMO profiles which closely mirrored those reported by a longitudinal study of US mothers followed over 24 months post-partum (35). Finally, the HMO profiles in mothers from north-eastern Uganda, who are predominantly of the Nilo-Hamitic Atesot tribe, may not be generalizable to mothers in other Ugandan regions with different environments, diets and tribes.

In conclusion, our studies of non-genetic determinants of HMO composition in rural Ugandan mothers identified specific HMOs which were associated with infant sex and maternal age, parity and BMI but were different from those reported in other populations, thereby underscoring the importance of generating population context-specific HMO data. Our study revealed HMO profiles in Ugandan mothers with prolonged lactation durations beyond 52 weeks which were remarkably similar to published HMO profiles from mothers with shorter lactation durations. The conservation of these HMO profiles in different populations, regardless of environments, diets and ethnicities, probably underscores their importance in infant/child growth and survival throughout lactation.

## Data Availability

Data described in the manuscript, code book, and analytic code will be made available upon request from TGE.

## Acknowledgments

We are very grateful to the mother-infant pairs for their generosity and participation. We would like to thank the clinical and laboratory staff of Med Biotech Laboratories at St Anne HC III Usuk, Katakwi, Uganda.

## Funding

Thomas Egwang received funding from the Global Innovation Fund and Grand Challenges Canada. Lars Bode is UC San Diego Chair of Collaborative Human Milk Research endowed by the Family Larsson-Rosenquist Foundation (FLRF), Switzerland. The funders played no role in the study design, data collection and analysis nor in the preparation of the manuscript and decision to publish this paper.

## Conflict of interest and funding disclosure

The authors have no conflicts of interest to disclose.

## Authors’ contributions to manuscript

TJO and EO conducted the study including sample collection; TJO drafted the manuscript; SM and CY performed the HMO laboratory analysis; VIM performed statistical analysis; TGE and LB conceived and directed the study. All authors read and approved the final manuscript.

## Abbreviations

2’FL: 2’fucosyllactose
3 FL: 3-fucosyllatose
3’SL: 3’sialyllactose
6’SL: 6’sialyllactose
BMI: body mass index
DFLac: difucosyllactose
DFLNH: Difucosyllacto-N-hexaose
DFLNT: Difucosyllacto-N-tetrose
DSLNH: disialyllacto-N-tetraose
DSLNT: Disialyllacto-N-tetraose
FDSLNH: Fucosyldisialyllacto-N-hexaose
FLNH: Fucosyllacto-N-hexaose
Fuc HMO: HMO-bound fucose
FUT2: α1,2-fucosyltransferase gene
HMOs: human milk oligosaccharides
LNFPI: lacto-N-fucopentaose I
LNFPII: lacto-N-fucopentaose II
LNFPIII: lacto-N-fucopentaose III
LNH: Lacto*-*N*-*hexaose
LNnT: Lacto-N-neotetraose
LNT: Lacto-N-tetraose
LSTb: Sialyllacto-*N*-tetraose b
LSTc: sialyllacto-N-tetraose c
Sia HMO: HMO-bound sialic acid

---

**Supplemental Figure 1.**
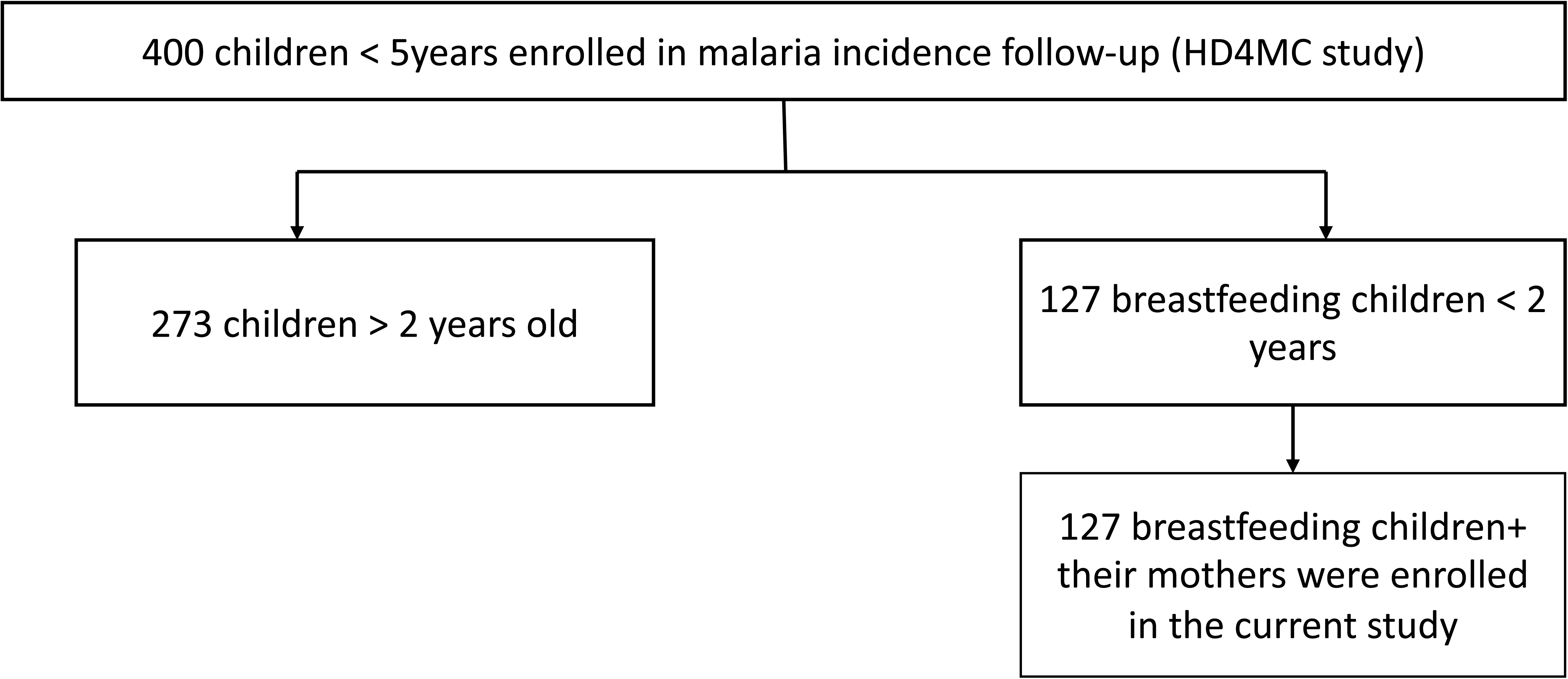
Study participants flow chart.

**Supplemental FIGURE 2.**
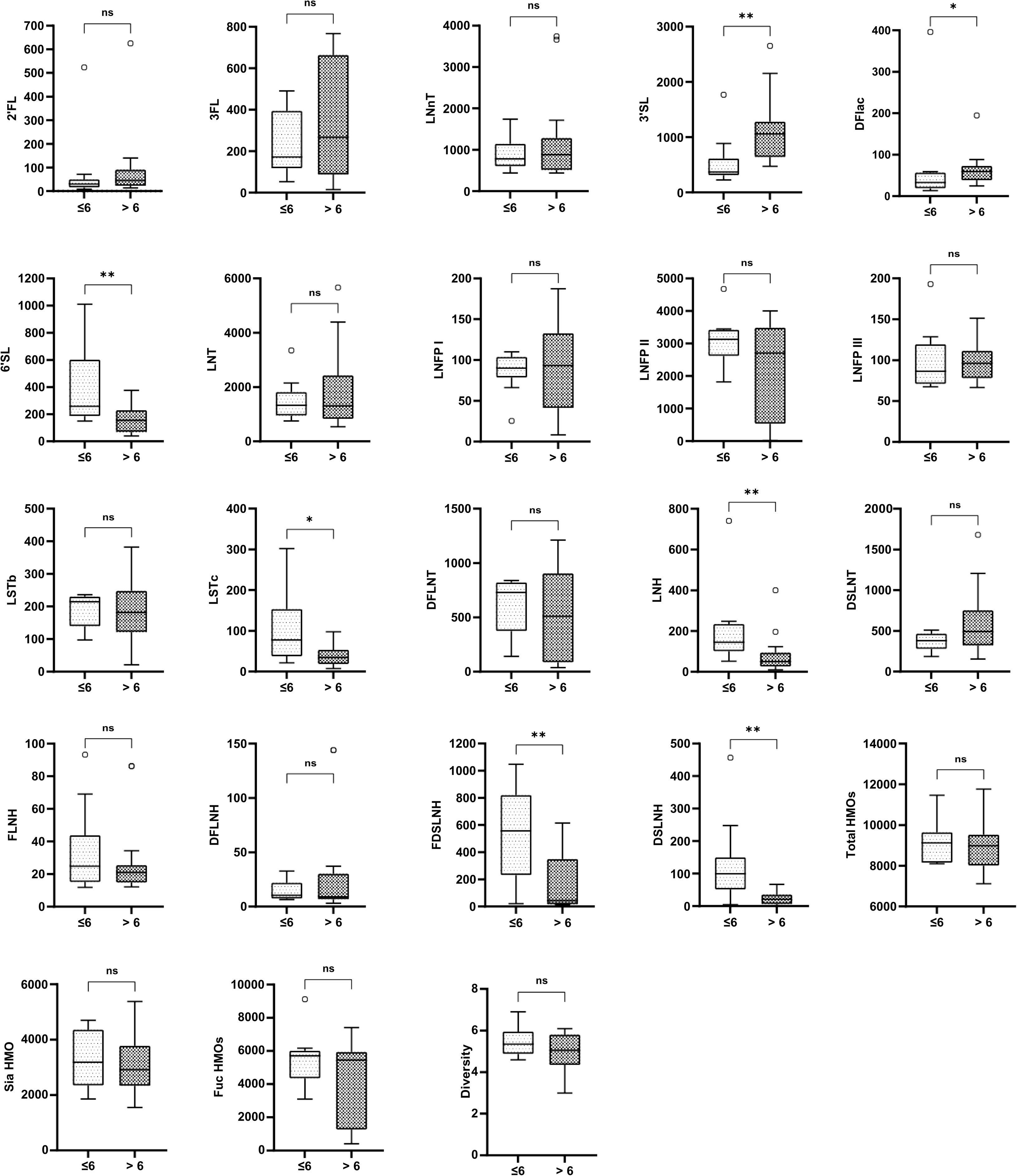
Concentrations of all measured HMOs before and after 6 months postpartum in breast milk from non-secretor Ugandan mother-child pairs. HMO name abbreviations are indicated in the list of abbreviations. Data are shown as box and whisker plots with the medians and the lower and upper quartiles or interquartile range (IQR). P values represent the outcomes of comparisons of two groups by the Mann-Whitney U test.

## Notes

### Competing Interest Statement

The authors have declared no competing interest.

