## Supplemental Tables for "The effect of non-genetic determinants of human milk oligosaccharide profiles in milk of Ugandan mothers"

Title: Non-genetic mother-infant factors and Human milk oligossacharides composition/profile in breast milk of Ugandan mothers

First Author: Tonny Jimmy Owalla

**Supplementary Table 1: Median and Percent HMO concentrations**

| | All | | Secretors | | Non-secretors | |
|---|---|---|---|---|---|---|
| **HMO** | **nmol/mL** | **%** | **nmol/mL** | **%** | **nmol/mL** | **%** |
| 2'FL | 5929.5 | 36.9 | 7099.7 | 42.5 | 31.7 | 0.3 |
| LNFP II | 1411.1 | 8.8 | 1140.5 | 6.8 | 2884.1 | 32.1 |
| LNT | 1028.7 | 6.4 | 981.3 | 5.8 | 1330.7 | 14.8 |
| DFLNT | 865.9 | 5.4 | 1215.9 | 7.3 | 619.0 | 6.9 |
| LNnT | 805.2 | 5.0 | 789.1 | 4.7 | 870.1 | 9.7 |
| 3'SL | 632.0 | 3.9 | 619.5 | 3.7 | 692.7 | 7.7 |
| LNFP I | 599.3 | 3.7 | 1019.8 | 6.1 | 91.5 | 1.0 |
| DFlac | 548.4 | 3.4 | 666.0 | 4.0 | 56.1 | 0.6 |
| DSLNT | 417.2 | 2.6 | 404.4 | 2.4 | 429.3 | 4.8 |
| 3FL | 378.9 | 2.4 | 397.7 | 2.4 | 189.3 | 2.1 |
| 6'SL | 157.8 | 1.0 | 150.9 | 0.9 | 185.9 | 2.1 |
| FDSLNH | 146.7 | 0.9 | 134.0 | 0.8 | 216.3 | 2.4 |
| LSTb | 123.5 | 0.8 | 119.2 | 0.7 | 183.8 | 2.0 |
| LNH | 71.2 | 0.4 | 68.6 | 0.4 | 80 | 0.9 |
| LNFP III | 70.0 | 0.4 | 63.5 | 0.4 | 93.8 | 1.0 |
| LSTc | 46.5 | 0.3 | 46.9 | 0.3 | 41.3 | 0.5 |
| DSLNH | 31.7 | 0.2 | 31.7 | 0.2 | 31.3 | 0.3 |
| DFLNH | 24.4 | 0.2 | 28.3 | 0.2 | 9.5 | 0.1 |
| FLNH | 24.2 | 0.2 | 25.8 | 0.2 | 21 | 0.2 |
| Total HMO | 16050.6 | | 16699.2 | | 8988.5 | 100 |
| Fuc HMO | 15962.3 | 99.5 | 14641.2 | 87.7 | 5611.3 | 62.4 |
| Sial HMO | 2395.0 | 14.9 | 2325.6 | 13.9 | 2984.1 | 33.2 |

HMOs = human milk oligosaccharides; nmol/mL = nanomoles/milliliter; % = percent

Title: Non-genetic mother-infant factors and Human milk oligossacharides composition/profile in breast milk of Ugandan mothers

First Author: Tonny Jimmy Owalla

**Supplementary Table 2: Median (IQR) HMO concentrations and sex of children**

| HMO nmol/mL | Total (N=114) | Females (n=60) | Males (n=54) | Unadjusted p-value |
|---|---|---|---|---|
| 2'FL | 5961.3 (2365.5-9364.7) | 5961.3 (3086.9-9616.9) | 6069.2 (1660.3-9271.8) | 0.97 |
| 3FL | 355.5 (263.8-593.5) | 371.2 (240.5-536.2) | 353.8 (289.4-660.3) | 0.35 |
| LNnT, | 782.3 (578.2-1042.6) | 768.5 (582.9-1037.4) | 813.1 (567.9-1093.1) | 0.65 |
| **3'SL** | **614.1 (352.5-964.9)** | **516.6 (309.8-853.8)** | **701.8 (454.5-1059.8)** | **0.035** |
| DFLac | 493.2 (208.5-832.6) | 457.6 (281.9-744.6) | 557.0 (194.8-961.2) | 0.57 |
| 6’SL | 154.0 (100.3-276.4) | 148.9 (101.3-230.4) | 183.0 (97.8-297.8) | 0.39 |
| LNT | 1010.4 (702.1-1618.0) | 982.2 (730.7-1513.2) | 1076.7 (659.6-1704.2) | 0.88 |
| LNFP I | 623.9 (169.2-1863.9) | 729.9 (224.4-2039.1) | 455.8 (169.2-1781.5) | 0.64 |
| LNFP II | 1393.9 (721.0-2582.7) | 1359.6 (759.1-2613.2) | 1476.8 (599.7-2529.6) | 0.73 |
| LNFP III | 71.0 (55.0-88.8) | 65.0 (54.2-84.6) | 75.0 (58.9-93.2) | 0.14 |
| LSTb | 123.3 (94.8-182.2) | 123.3 (101.3-178.7) | 124.7 (83.3-182.2) | 0.62 |
| LSTc, | 43.1 (22.2-92.4) | 41.8 (23.4-72.3) | 45.8 (21.4-104.3) | 0.71 |
| DFLNT | 837.2 (106.0-1609.2) | 837.2 (129.8-1666.7) | 868.0 (80.4-1544.0) | 0.97 |
| LNH | 67.8 (37.2-107.4) | 74.8 (41.6-125.0) | 60.7 (30.7-82.9) | 0.16 |

| | | | | |
|---|---|---|---|---|
| DSLNT | 425.8 (280.5-515.5) | 377.7 (280.6-476.5) | 443.6 (274.3-585.6) | 0.12 |
| FLNH | 24.0 (13.2-52.0) | 25.3 (13.1-50.9) | 23.7 (13.2-58.5) | 0.86 |
| DFLNH | 25.6 (10.1-66.1) | 26.6 (10.5-66.1) | 25.1 (9.3-66.1) | 0.86 |
| FDSLNH | 144.7 (21.1-290.3) | 171.5 (16.9-313.6) | 102.2 (23.1-237.9) | 0.54 |
| DSLNH | 26.1 (11.1-63.7) | 24.3 (7.8-81.0) | 30.9 (12.0-62.5) | 0.95 |
| Total HMO | 16021.3 (12674.9-17668.9) | 15804.3 (12947.1-17249.5) | 16174.0 (12197.8-18096.5) | 0.43 |
| Sia HMO | 13985.2 (10010.4-15618.4) | 13756.7 (9913.9-15226.6) | 14186.8 (10517.4-16252.6) | 0.49 |
| Fuc HMO | 2388.4 (1965.7-3032.4) | 2322.1 (1836.1-2861.0) | 2403.0 (2026.4-3305.9) | 0.17 |
| Diversity | 4.7 (3.0-5.9) | 4.7 (3.0-6.0) | 4.7 (3.0-5.9) | 0.96 |

IQR = interquartile range; HMOs = human milk oligosaccharides; N = total number of mothers with available information about the sex of their children; data on sex was not available for 13 children; n = number of females or males. HMO with significant unadjusted p value is in bold.

Title: Non-genetic mother-infant factors and Human milk oligossacharides composition/profile in breast milk of Ugandan mothers

First Author: Tonny Jimmy Owalla

**Supplementary Table 3: Correlations between HMO concentrations and lactation duration**

| HMOs nmol/mL | All, N= 127 | | Secretor, n=101 | | Non Secretor, n= 26 | |
|---|---|---|---|---|---|---|
| | Coef. (rho) | Unadjusted p-value | Coef. (rho) | Unadjusted p-value | Coef. (rho) | Unadjusted p-value |
| 2'FL | 0.0479 | 0.5925 | 0.0729 | 0.4686 | 0.2652 | 0.1904 |
| **3FL** | **0.3551** | **<0.0001** | **0.3626** | **0.0002** | 0.3802 | 0.0554 |
| **LNnT,** | **-0.3680** | **<0.0001** | **-0.4188** | **<0.0001** | -0.2440 | 0.2297 |
| **3'SL** | **0.4104** | **<0.0001** | **0.3378** | **0.0006** | **0.7062** | **0.0001** |
| **DFLac** | **0.2740** | **0.0018** | **0.4214** | **<0.0001** | **0.5195** | **0.0065** |
| **6'SL** | **-0.6915** | **<0.0001** | **-0.6586** | **<0.0001** | **-0.8010** | **<0.0001** |
| **LNT** | **-0.1993** | **0.0247** | -0.1919 | 0.0545 | -0.2785 | 0.1683 |
| LNFP I | **-0.2133** | **0.0160** | **-0.3105** | **0.0016** | 0.0544 | 0.7918 |
| LNFP II | 0.1097 | 0.2194 | **0.2002** | **0.0448** | -0.0400 | 0.8460 |
| LNFP III | -0.0596 | 0.5056 | -0.0545 | 0.5881 | -0.0127 | 0.9510 |
| LSTb | 0.0087 | 0.9226 | -0.0250 | 0.8043 | 0.1417 | 0.4900 |
| **LSTc** | **-0.6630** | **<0.0001** | **-0.6429** | **<0.0001** | **-0.7343** | **<0.0001** |
| DFLNT | -0.1125 | 0.2080 | -0.1406 | 0.1607 | 0.1649 | 0.4207 |
| **LNH** | **-0.2756** | **0.0017** | **-0.2100** | **0.0350** | **-0.4660** | **0.0164** |
| DSLNT | -0.0346 | 0.6994 | -0.1501 | 0.1340 | **0.3904** | **0.0486** |
| **FLNH** | **-0.3090** | **0.0004** | **-0.3334** | **0.0007** | -0.1393 | 0.4975 |
| **DFLNH** | **-0.2411** | **0.0063** | **-0.2860** | **0.0037** | -0.0339 | 0.8694 |
| FDSLNH | -0.0427 | 0.6333 | 0.0994 | 0.3226 | **-0.4722** | **0.0149** |
| **DSLNH** | **-0.5310** | **<0.0001** | **-0.5026** | **<0.0001** | **-0.6223** | **0.0007** |
| Total HMO | -0.0183 | 0.8383 | 0.0070 | 0.9450 | -0.1718 | 0.4014 |
| Fuc HMO | 0.0937 | 0.2948 | 0.1691 | 0.0909 | 0.0366 | 0.8591 |
| Sia HMO | -0.0631 | 0.4806 | -0.0709 | 0.4811 | -0.0106 | 0.9590 |
| Diversity | -0.0989 | 0.2687 | -0.1030 | 0.3056 | -0.1271 | 0.5362 |

Coef (rho) = Spearman's correlation coefficient. HMOs = human milk oligosaccharides; nmol/mL = nanomoles/milliliter; N = total number of mothers; n = number of secretors or non-secretors. HMOs with significant unadjusted p value are in bold.

Title: Non-genetic mother-infant factors and Human milk oligossacharides composition/profile in breast milk of Ugandan mothers

First Author: Tonny Jimmy Owalla

**Supplementary Table 4: Correlations between HMO concentrations and maternal age**

| HMOs nmol/mL | All, N= 126 | | Secretor, N= 100 | | Non-Secretor, N= 26 | |
|---|---|---|---|---|---|---|
| | **Coef. (rho)** | **Unadjusted p-value** | **Coef. (rho)** | **Unadjusted p-value** | **Coef. (rho)** | **Unadjusted p-value** |
| **2'FL** | 0.0410 | 0.6488 | 0.1228 | 0.2235 | **0.4584** | **0.0185** |
| 3FL | 0.0737 | 0.4124 | 0.1500 | 0.1363 | -0.0646 | 0.7539 |
| **LNnT** | **-0.2213** | **0.0128** | **-0.3040** | **0.0021** | 0.0357 | 0.8624 |
| 3'SL | 0.1753 | 0.0505 | 0.1302 | 0.1989 | 0.3591 | 0.0716 |
| **DFLac** | 0.0537 | 0.5505 | 0.1403 | 0.1639 | **0.4506** | **0.0209** |
| 6’SL | -0.0344 | 0.7024 | -0.0409 | 0.6862 | -0.0351 | 0.8650 |
| LNT | -0.1152 | 0.1989 | -0.1559 | 0.1214 | -0.0213 | 0.9177 |
| LNFP I | -0.0767 | 0.3933 | -0.0103 | 0.9192 | -0.3560 | 0.0742 |
| LNFP II | -0.0080 | 0.9288 | 0.0090 | 0.9292 | -0.1625 | 0.4276 |
| LNFP III | 0.0556 | 0.5362 | 0.0313 | 0.7569 | -0.0653 | 0.7513 |
| LSTb | -0.0153 | 0.8651 | -0.0632 | 0.5324 | 0.2285 | 0.2615 |
| LSTc | -0.0684 | 0.4468 | -0.0925 | 0.3602 | 0.0502 | 0.8077 |
| DFLNT | -0.1103 | 0.2188 | -0.0844 | 0.4038 | -0.0443 | 0.8297 |
| LNH | -0.1639 | 0.0667 | -0.1699 | 0.0911 | -0.1966 | 0.3358 |
| **DSLNT** | -0.0673 | 0.4542 | **-0.2181** | **0.0293** | **0.4612** | **0.0177** |
| FLNH | -0.0372 | 0.6790 | -0.0628 | 0.5348 | 0.1887 | 0.3560 |
| DFLNH | 0.0895 | 0.3190 | 0.1379 | 0.1713 | 0.0160 | 0.9382 |
| FDSLNH | 0.0531 | 0.5551 | 0.0798 | 0.4300 | -0.1378 | 0.5020 |
| DSLNH | -0.0254 | 0.7779 | -0.0335 | 0.7411 | 0.0184 | 0.9290 |
| Total HMO | 0.0343 | 0.7030 | 0.1344 | 0.1826 | 0.0818 | 0.6912 |
| **Fuc HMO** | 0.0957 | 0.2863 | **0.2423** | **0.0151** | -0.0704 | 0.7324 |
| Sia HMO | -0.0093 | 0.9177 | -0.1060 | 0.2941 | 0.2598 | 0.1999 |
| Diversity | -0.1091 | 0.2240 | -0.1425 | 0.1572 | -0.0637 | 0.7571 |

Coef (rho) = Spearman’s correlation coefficient. HMOs = human milk oligosaccharides; nmol/mL = nanomoles/milliliter; N = total number of mothers; n = number of secretors or non-secretors. HMOs with significant unadjusted p value are in bold.

Title: Non-genetic mother-infant factors and Human milk oligossacharides composition/profile in breast milk of Ugandan mothers

First Author: Tonny Jimmy Owalla

**Supplementary Table 5: Median (IQR) HMO concentrations and maternal age**

| HMOs nmol/mL | Total (N=127) | >18 years (n=123) | ≤ 18years (n=4) | Unadjusted p-value |
|---|---|---|---|---|
| 2'FL | 5912.6 (2715.8-9271.8) | 5822.8 (2452.3-9279.6) | 7181.2 (5115.1-8249.1) | 0.73 |
| 3FL | 375.4 (263.8-593.5) | 384.4 (263.8-606.8) | 324.9 (257.7-380.7) | 0.43 |
| LNnT | 797.2 (578.2-1033.2) | 775.1 (567.9-1033.2) | 966.2 (872.0-1497.4) | 0.12 |
| 3'SL | 630.0 (353.4-985.9) | 640.2 (364.9-987.8) | 434.3 (303.9-545.7) | 0.18 |
| DFLac | 546.5 (287.0-838.1) | 546.5 (231.6-881.5) | 582.7 (385.4-722.3) | 0.92 |
| 6’SL | 158.5 (102.5-270.9) | 156.6 (102.5-263.3) | 480.3 (272.6-909.2) | 0.10 |
| LNT | 1039.5 (702.1-1618.0) | 1039.5 (694.8-1531.9) | 1267.2 (834.4-2033.7) | 0.48 |
| **LNFP I** | **609.7 (172.6-1863.9)** | **579.4 (169.2-1846.9)** | **2279.7 (1137.6-3344.6)** | **0.050** |
| LNFP II | 1393.9 (721.0-2551.2) | 1419.6 (735.3-2551.2) | 360.8 (147.4-1569.8) | 0.23 |
| LNFP III | 70.5 (55.4-88.8) | 70.5 (57.1-89.8) | 60.3 (45.2-74.1) | 0.28 |
| LSTb | 123.3 (90.8-182.2) | 123.2 (89.1-181.9) | 202.0 (113.5-291.4) | 0.19 |
| LSTc | 46.6 (24.8-93.3) | 45.7 (24.8-87.9) | 201.3 (70.3-690.2) | 0.11 |
| DFLNT | 852.8 (122.3-1544.0) | 852.8 (122.3-1526.3) | 1085.4 (190.7-1916.3) | 0.68 |
| LNH | 70.7 (37.7-105.2) | 71.5 (38.2-106.4) | 28.5 (24.6-61.9) | 0.16 |

| | | | | |
|---|---|---|---|---|
| **DSLNT** | **412.1 (274.3-512.6)** | **400.9 (271.0-504.0)** | **641.2 (600.7-922.3)** | **0.009** |
| FLNH | 24.2 (13.2-52.0) | 24.0 (13.2-52.0) | 28.3 (17.7-70.8) | 0.80 |
| DFLNH | 24.5 (9.4-66.1) | 23.5 (9.3-66.1) | 34.3 (26.9-100.3) | 0.24 |
| FDSLNH | 146.9 (21.1-310.1) | 157.5 (21.2-311.1) | 31.3 (14.9-106.7) | 0.16 |
| DSLNH | 32.1 (11.6-64.5) | 32.1 (11.6-64.5) | 35.5 (15.3-219.8) | 0.68 |
| Total HMO | 16021.3 (13219.4-17668.9) | 15907.3 (13190.4-17550.4) | 17502.3 (15383.9-18768.4) | 0.22 |
| Sia HMO | 13925.6 (10903.9-15947.2) | 13925.6 (10517.4-15947.2) | 13607.5 (11691.6-15523.7) | 0.97 |
| Fuc HMO | 2388.4 (1995.9-3149.7) | 2381.0 (1977.8-3114.2) | 3202.3 (2500.3-4456.0) | 0.10 |
| Diversity | 4.7 (3.1-6.0) | 4.7 (3.0-6.0) | 4.8 (3.6-6.5) | 0.52 |

IQR = interquartile range; HMOs = human milk oligosaccharides; N = total number of mothers; n = number of mothers in each age category. HMOs with significant unadjusted p values are in bold.

Title: Non-genetic mother-infant factors and Human milk oligossacharides composition/profile in breast milk of Ugandan mothers

First Author: Tonny Jimmy Owalla

**Supplementary Table 6: Median (IQR) HMO concentrations and maternal parity**

| HMOs nmol/mL | Total (N=127) | Parity=1 (n=27) | Parity >1 (n=100) | Unadjusted p-value |
|---|---|---|---|---|
| 2'FL | 5929.5 (2715.8-9271.8) | 6242.8 (624.9-8908.1) | 5912.6 (3799.2-9275.7) | 0.56 |
| 3FL | 378.9 (263.8-606.8) | 327.1 (221.0-406.1) | 401.0 (265.2-632.7) | 0.076 |
| LNnT | 805.2 (578.2-1033.5) | 894.2 (594.1-1141.3) | 790.0 (573.0-1032.7) | 0.28 |
| 3'SL | 632.0 (353.4-987.8) | 413.1 (295.2-985.9) | 700.6 (403.4-1016.9) | 0.072 |
| **DFLac** | **548.4 (287.0-881.5)** | **399.3 (158.4-698.6)** | **594.8 (365.0-937.9)** | **0.018** |
| 6'SL | 157.8 (102.2-270.9) | 182.2 (86.3-569.7) | 155.1 (102.8-253.6) | 0.38 |
| LNT | 1028.7 (694.8-1618.0) | 1106.3 (752.7-1747.0) | 1026.5 (666.2-1473.4) | 0.65 |
| LNFP I | 599.3 (172.6-1863.9) | 1080.8 (117.9-2066.2) | 529.4 (212.1-1730.5) | 0.44 |
| LNFP II | 1411.1 (721.0-2551.2) | 1580.5 (107.9-2766.3) | 1393.9 (787.8-2495.3) | 0.77 |
| LNFP III | 70.0 (55.0-88.8) | 68.4 (44.9-82.5) | 73.9 (58.8-93.2) | 0.11 |
| LSTb | 123.5 (90.8-182.2) | 134.5 (87.6-184.4) | 122.3 (92.8-181.8) | 0.51 |
| LSTc | 46.5 (24.3-93.3) | 63.7 (18.5-140.8) | 43.1 (25.2-75.1) | 0.19 |
| DFLNT | 865.9 (122.3-1576.6) | 802.1 (121.3-1705.9) | 993.8 (126.1-1560.3) | 0.91 |
| LNH | 71.2 (37.7-106.4) | 68.6 (23.0-104.0) | 71.8 (41.6-107.8) | 0.40 |
| DSLNT | 417.2 (274.3-512.6) | 454.3 (287.3-633.3) | 412.1 (268.7-501.5) | 0.30 |

| | | | | |
|---|---|---|---|---|
| FLNH | 24.2 (13.2-57.0) | 20.5 (8.3-35.6) | 24.5 (13.6-59.5) | 0.23 |
| DFLNH | 24.4 (9.4-66.1) | 16.9 (9.0-32.8) | 26.1 (10.1-74.3) | 0.21 |
| FDSLNH | 146.7 (21.1-310.1) | 123.9 (15.8-337.3) | 157.7 (25.5-302.0) | 0.39 |
| DSLNH | 31.7 (11.6-64.5) | 30.7 (6.5-65.8) | 32.1 (12.7-64.1) | 0.98 |
| Total HMO | 16050.6 (13219.4-17668.9) | 15755.6 (10981.1-17187.8) | 16125.7 (13379.3-17682.1) | 0.73 |
| Sia HMO | 13962.3 (10903.9-15966.1) | 13381.6 (7401.8-15258.1) | 14117.4 (11166.4-16134.6) | 0.22 |
| Fuc HMO | 2395.0 (1995.9-3149.7) | 2398.6 (1795.0-3301.5) | 2388.4 (1995.9-3022.9) | 0.76 |
| Diversity | 4.7 (3.1-6.0) | 4.3 (3.1-6.0) | 4.7 (3.1-6.0) | 0.91 |

IQR = interquartile range; HMOs = human milk oligosaccharides; N = total number of mothers; n = number of mothers in each parity category. HMO with a significant unadjusted p value is in bold.

Title: Non-genetic mother-infant factors and Human milk oligossacharides composition/profile in breast milk of Ugandan mothers

First Author: Tonny Jimmy Owalla

**Supplementary Table 7: Correlations between HMOs and maternal post-partum BMI**

| HMO nmol/mL | All, N= 113 | | Secretor, n= 89 | | Non-Secretor, n= 24 | |
|---|---|---|---|---|---|---|
| | Coef. (rho) | Unadjusted p-value | Coef. (rho) | Unadjusted p-value | Coef. (rho) | Unadjusted p-value |
| 2'FL | -0.0547 | 0.5651 | -0.0662 | 0.5376 | -0.1283 | 0.5502 |
| 3FL | 0.0184 | 0.8469 | 0.0502 | 0.6401 | -0.1313 | 0.5407 |
| LNnT | -0.0266 | 0.7794 | -0.0219 | 0.8386 | -0.0565 | 0.7930 |
| 3'SL | 0.1361 | 0.1525 | 0.1700 | 0.1134 | -0.0291 | 0.8925 |
| DFLac | 0.0255 | 0.7888 | 0.0580 | 0.5890 | -0.0726 | 0.7359 |
| 6’SL | 0.0088 | 0.9259 | -0.0310 | 0.7733 | 0.2392 | 0.2603 |
| LNT | 0.0521 | 0.5837 | 0.0123 | 0.9092 | 0.1913 | 0.3704 |
| LNFP I | -0.0848 | 0.3717 | -0.1091 | 0.3090 | -0.0939 | 0.6624 |
| LNFP II | 0.0703 | 0.4594 | 0.1374 | 0.1992 | -0.1366 | 0.5246 |
| LNFP III | 0.0667 | 0.4824 | 0.0805 | 0.4534 | -0.0992 | 0.6448 |
| LSTb | -0.0196 | 0.8364 | 0.0308 | 0.7745 | -0.1992 | 0.3508 |
| LSTc | -0.0279 | 0.7695 | -0.0249 | 0.8171 | -0.0918 | 0.6698 |
| DFLNT | -0.0965 | 0.3093 | -0.1180 | 0.2709 | -0.2231 | 0.2947 |
| **LNH** | -0.1641 | 0.0824 | -0.0917 | 0.3926 | **-0.4188** | **0.0417** |
| DSLNT | 0.0017 | 0.9855 | -0.0297 | 0.7824 | 0.0952 | 0.6580 |
| FLNH | -0.0383 | 0.6868 | -0.0084 | 0.9380 | -0.2092 | 0.3266 |
| DFLNH | -0.0217 | 0.8194 | -0.0066 | 0.9512 | -0.1653 | 0.4401 |
| **FDSLNH** | 0.1107 | 0.2431 | **0.2126** | **0.0454** | -0.2405 | 0.2577 |
| DSLNH | -0.0712 | 0.4538 | -0.0718 | 0.5035 | -0.1131 | 0.5988 |
| Total HMO | -0.0234 | 0.8059 | -0.0108 | 0.9200 | -0.2592 | 0.2213 |
| Fuc HMO | -0.0649 | 0.4944 | -0.0946 | 0.3777 | -0.0822 | 0.7026 |
| Sia HMO | 0.0718 | 0.4500 | 0.1272 | 0.2348 | -0.2283 | 0.2833 |
| Diversity | 0.0311 | 0.7439 | 0.0585 | 0.5860 | -0.1864 | 0.3833 |

Coef (rho) = Spearman’s correlation coefficient. HMO = human milk oligosaccharides; nmol/mL = nanomoles/milliliter; N = total number of mothers with BMI data available; height or weight data were unavailable for 14 mothers; n = number of secretors or non-secretors. HMOs with significant unadjusted p values are in bold.

Title: Non-genetic mother-infant factors and Human milk oligossacharides composition/profile in breast milk of Ugandan mothers

First Author: Tonny Jimmy Owalla

**Supplementary Table 8: Median (IQR) HMO concentrations and Maternal post-partum BMI in Secretor Mothers**

| HMO nmol/mL | Total (N=101) | Normal (n=82) | Overweight (n=19) | Unadjusted p-value |
|---|---|---|---|---|
| 2'FL | 7059.5 (4978.3-9808.0) | 7215.0 (4991.9-9774.8) | 6242.8 (4468.4-10037.1) | 0.43 |
| 3FL | 399.2 (294.5-632.7) | 400.8 (297.8-633.6) | 370.5 (212.5-543.2) | 0.32 |
| LNnT | 797.2 (558.8-1033.4) | 805.2 (594.1-1033.5) | 774.7 (439.5-1026.8) | 0.46 |
| 3'SL | 624.8 (346.2-916.9) | 630.0 (364.9-916.3) | 528.1 (309.0-958.8) | 0.71 |
| DFLac | 658.6 (451.6-1041.7) | 698.6 (457.1-1128.3) | 480.8 (404.8-888.2) | 0.28 |
| 6’SL | 150.7 (99.0-270.9) | 150.5 (97.8-263.3) | 150.9 (107.7-320.8) | 0.70 |
| LNT | 989.0 (649.9-1473.4) | 964.4 (653.7-1395.3) | 1106.3 (485.9-1625.3) | 0.78 |
| LNFP I | 974.5 (387.9-2112.2) | 845.0 (382.9-2090.4) | 1230.9 (537.4-2133.9) | 0.80 |
| LNFP II | 1154.3 (558.1-2089.4) | 1106.0 (535.5-2085.7) | 1580.5 (897.6-2273.7) | 0.22 |
| LNFP III | 63.3 (49.3-81.6) | 63.5 (52.4-81.2) | 61.7 (45.0-88.0) | 0.73 |
| LSTb | 119.4 (85.6-159.0) | 119.2 (87.4-142.8) | 126.4 (65.7-216.6) | 0.61 |
| LSTc | 46.8 (23.9-95.2) | 46.7 (23.3-77.2) | 58.3 (31.5-137.3) | 0.31 |
| DFLNT | 1223.6 (128.0-1739.9) | 1313.1 (190.2-1815.0) | 144.8 (78.7-1421.5) | 0.14 |
| LNH | 69.4 (37.6-97.9) | 73.2 (37.2-103.1) | 51.3 (41.2-83.7) | 0.38 |

| | | | | |
|---|---|---|---|---|
| DSLNT | 405.7 (261.4-505.7) | 426.3 (263.2-512.6) | 320.0 (260.2-456.4) | 0.21 |
| FLNH | 26.0 (11.8-58.5) | 26.2 (11.9-58.5) | 15.8 (9.1-42.0) | 0.44 |
| DFLNH | 28.1 (12.1-74.3) | 28.0 (11.5-67.4) | 28.3 (16.7-76.6) | 0.86 |
| **FDSLNH** | **140.3 (20.8-270.6)** | **102.2 (18.3-237.9)** | **213.6 (107.1-337.3)** | **0.028** |
| DSLNH | 31.4 (12.7-61.8) | 29.1 (11.6-62.1) | 33.7 (15.8-60.0) | 0.62 |
| Total HMO | 16650.0 (15344.7-17878.3) | 16725.9 (15434.2-18096.5) | 16237.7 (14883.9-17695.3) | 0.35 |
| Sia HMO | 2329.0 (1930.1-2836.4) | 2335.1 (1934.2-2796.0) | 2269.9 (1760.5-3169.8) | 0.92 |
| Fuc HMO | 14638.9 (13461.9-16231.6) | 14641.2 (13750.5-16277.2) | 13528.5 (11826.2-16182.3) | 0.086 |
| Diversity | 4.4 (2.9-6.1) | 4.1 (2.9-6.2) | 5.0 (2.8-6.0) | 0.77 |

IQR = interquartile range; HMOs = human milk oligosaccharides; N = total number of mothers; n = number of mothers in each weight category. HMO with a significant unadjusted p value is in bold.

Title: Non-genetic mother-infant factors and Human milk oligossacharides composition/profile in breast milk of Ugandan mothers

First Author: Tonny Jimmy Owalla

**Supplementary Table 9: Median (IQR) HMO concentrations and Maternal BMI in Non-secretor Mothers**

| HMO nmol/mL | Total (N=26) | Normal (n=20) | Overweight (n=6) | Unadjusted p-value |
|---|---|---|---|---|
| 2'FL | 31.7 (21.0-69.8) | 35.7 (23.5-67.6) | 27.2 (17.8-98.0) | 0.81 |
| 3FL | 189.3 (130.8-515.8) | 235.6 (104.6-553.2) | 185.1 (178.6-378.9) | 0.81 |
| LNnT | 870.1 (585.6-1203.5) | 870.1 (587.4-1257.5) | 760.0 (585.6-914.7) | 0.67 |
| 3'SL | 692.7 (474.1-1091.4) | 692.7 (430.9-1101.7) | 850.0 (601.5-1059.8) | 0.86 |
| DFLac | 56.1 (31.1-65.1) | 53.4 (30.0-69.0) | 59.2 (41.3-61.4) | 0.63 |
| 6’SL | 185.9 (149.7-297.8) | 185.9 (149.8-336.9) | 200.9 (65.9-243.9) | 0.67 |
| LNT | 1330.7 (905.1-2293.9) | 1330.7 (841.1-1986.6) | 1704.9 (964.6-2426.7) | 0.47 |
| LNFP I | 91.5 (54.4-105.9) | 90.1 (49.2-108.0) | 97.5 (82.7-102.1) | 0.90 |
| LNFP II | 2884.1 (1836.4-3417.2) | 2884.1 (2356.9-3360.9) | 2626.8 (1836.4-3507.9) | 0.95 |
| LNFP III | 93.8 (76.6-111.6) | 89.9 (74.3-114.0) | 98.0 (86.5-101.5) | 0.81 |
| LSTb | 183.8 (123.1-230.0) | 205.1 (150.6-231.2) | 131.2 (21.4-185.3) | 0.11 |
| LSTc | 41.3 (26.7-67.1) | 41.3 (26.8-77.5) | 48.1 (20.0-57.5) | 0.81 |
| DFLNT | 619.0 (121.3-839.7) | 663.6 (197.8-852.8) | 341.1 (80.4-834.7) | 0.43 |
| **LNH** | **80.0 (37.7-155.1)** | **109.7 (60.5-179.2)** | **36.4 (26.5-54.4)** | **0.024** |
| DSLNT | 429.3 (315.3-593.0) | 425.9 (284.1-610.7) | 432.2 (346.2-593.0) | 0.76 |
| FLNH | 21.0 (15.2-34.2) | 21.0 (16.5-32.1) | 19.5 (15.1-86.2) | 0.86 |

| | | | | |
|---|---|---|---|---|
| DFLNH | 9.5 (7.7-22.8) | 10.1 (7.8-21.1) | 8.3 (6.7-144.1) | 0.95 |
| FDSLNH | 216.3 (21.2-554.2) | 338.5 (21.1-588.0) | 90.5 (38.4-247.4) | 0.36 |
| DSLNH | 31.3 (7.9-77.4) | 31.6 (7.9-81.8) | 29.4 (6.8-35.7) | 0.50 |
| Total HMO | 8988.5 (8159.9-9535.5) | 9246.0 (8373.8-9568.8) | 8450.6 (7941.6-8965.8) | 0.13 |
| Sia HMO | 2984.1 (2380.1-3986.7) | 3132.0 (2631.3-4261.7) | 2395.0 (2311.8-2901.2) | 0.10 |
| Fuc HMO | 5611.3 (3099.0-5958.7) | 5611.3 (4120.2-5922.5) | 4422.9 (3009.3-5966.2) | 0.76 |
| Diversity | 5.3 (4.6-5.9) | 5.3 (4.9-5.9) | 4.6 (4.6-5.4) | 0.13 |

IQR = interquartile range; HMOs = human milk oligosaccharides; N = total number of mothers; n = number of mothers in each weight category. HMO with a significant unadjusted p value is in bold.
